# Knockdown of PAK1IP1 can induce CASP-3-dependent pyroptosis to inhibit the progression of hepatocellular carcinoma

**DOI:** 10.1101/2024.06.02.24308341

**Authors:** Xiaoliang Lu, Jie Chen, Zefa Lu, Hong Zang

## Abstract

**Background:** Hepatocellular carcinoma (HCC), is a prevalent and fatal malignancy originating from hepatic cells with a consistently rising incidence in recent decades. In this study, we aim to identify potential prognostic biomarkers and reveal new mechanism in HCC.

**Methods:** HCC-related datasets (GSE45267 and GSE49515) and TCGA information were downloaded for DEGs, and the common DEGs were WGCNA, protein-protein interaction network (PPI), risk model, expression, survival and prognostic nomogram to determine the key gene related to HCC. Further, the key gene was analyzed by clinical feature analysis, immunoassay and cell experiments to investigate its exact role in HCC.

**Results:** Based on the above comprehensive analysis, we targeted the key gene PAK1-interacting protein 1(PAK1IP1) with a good prognostic value in HCC. PAK1IP1 was remarkably increased in tumor samples than normal samples, which might be related to immune cell infiltration in liver cancer. It was up-regulated in HCC cells, and its knockdown could suppress HCC proliferation and migration. Besides, ELISA and flow cytometry showed that PAK1IP1 could regulate Lipopolysaccharide (LPS)-induced pyroptosis of HCC cells. Knocking down PAK1IP1 could induce CASP-3-dependent pyroptosis in HCC cells to suppress the development of HCC.

**Conclusion:** To sum up, PAK1IP1 was identified as a promising prognostic biomarker, and knockdown of PAK1IP1 can induce CASP-3-dependent pyroptosis to suppress HCC development, which sheds new light on HCC tumorigenesis.

## Introduction

Liver cancer, or hepatocellular carcinoma (HCC), is a prevalent and fatal malignancy originating from hepatic cells[1]. Its incidence has shown a consistent rise in recent decades, making it one among the most common malignancies to be diagnosed globally[2]. The current treatment options for HCC encompass surgical intervention, chemotherapy, radiation therapy, targeted therapy, and immunotherapy[3]. The appropriate treatment modality is up to cancer stage, overall health status, liver function and other factors[4]. In spite of the availability of several therapeutic options, the 5-year survival rate for individuals with HCC is still poor, at about 18%. Thus, the development of a reliable prognostic model that integrates clinical, laboratory, and molecular biomarkers is crucial for guiding the clinical management of HCC patients[5]. Prognostic factors commonly employed in HCC encompass clinical parameters, laboratory data, radiological findings, and molecular biomarkers[6]. With the advancement of artificial intelligence and machine learning, numerous studies have focused on constructing predictive models to enhance the accuracy of prognostic predictions, which holds great promise in providing valuable insights for the customized therapy of HCC patients.

Pyroptosis was initially discovered in the 1970s[7]. It is a form of programmed cell death (PCD) characterized by cellular swelling, culminating in cell membrane rupture and the release of cellular contents, thereby inciting a robust inflammatory response[8]. This process of inflammatory cell death, also known as cellular inflammatory necrosis, plays a pivotal role in tumor suppression by stimulating anti-tumor immune responses. Pyroptosis is orchestrated by the activation of the inflammasome upon sensing endogenous danger signals or environmental stimuli, which recruits and activates caspase[9]. Activated caspase has a dual role; it not only cleaves and activates inflammatory factors such as IL-18 and IL-1β but also cleaves Gasdermin-D (GSDMD), inducing cell membrane perforation and triggering cell pyroptosis[10]. In the context of liver cancer, researchers have found that inducing pyroptosis holds significant therapeutic potential for cancer treatment. Several studies have demonstrated that liver cancer cells are more susceptible to pyroptosis induction compared to normal cells[11, 12]. Furthermore, the regulation of HCC cell death through pyroptosis involves various factors, including the Bcl-2 protein family and the tumor suppressor gene p53[13]. Pyroptosis is a well-established cellular mechanism of cell death that has drawn interest from cancer researchers because of its potential applications in cancer therapy[14]. The mechanism and potential uses of copper-dependent pyroptosis in inducing cancer cell death are still under exploration. In liver cancer, inducing pyroptosis shows great promise as a therapeutic approach, and ongoing research in this field aims to enhance our understanding of cancer biology, particularly in HCC cells.

In this comprehensive study, we employed a combination of computational and experimental approaches to investigate HCC. Initially, we curated and analyzed datasets from Gene expression omnibus (GEO) and the cancer genome atlas (TCGA), utilizing bioinformatics tools to determine key prognostic genes related to HCC. Subsequently, we developed a prognostic risk model and prognostic nomogram to evaluate gene clinical significance. In addition, we employed thorough analyses of clinical features and immune profiles to explore a comprehensive knowledge of these key genes in liver cancer. Subsequently, we conducted cell experiments to investigate the mechanisms by which these key genes, as well as pyroptosis-related genes, regulate the development of HCC. Overall, this research contributed new understanding of HCC and offered potential targets for clinical applications, offering promising avenues for improved diagnosis and treatment strategies in the field of HCC.

## Materials and methods

### Data origination

We obtained 371 Liver hepatocellular carcinoma (LIHC) samples and 50 normal samples from TCGA. Additionally, we downloaded two publicly available datasets GSE45267 and GSE49515 from GEO. Raw data from TCGA and GEO were preprocessed using the “affy” and “limma” packages in R software. Clinical information of patients, including age, sex, stage, and survival data, was extracted from the TCGA database.

### Weighted gene co-expression network analysis (WGCNA)

Differentially expressed genes (DEGs) identification was screened on three datasets: GSE45267, GSE49515 and TCGA. Venn diagrams were used to identify overlapping up- and down-regulated genes. WGCNA is a powerful bioinformatics tool that can identify groups of highly related genes and their relationship to phenotypic traits. We utilized WGCNA to analyze the overlapping genes to determine key module for the following analysis.

### Functional enrichment analysis and MCODE analysis

Next, Gene Ontology (GO) and Kyoto Encyclopedia of Genes and Genomes (KEGG) analysis were conducted. *P*<0.05 was used to determine the significance of the enriched results. Additionally, the co-expression network of important genes had major modules that we were able to locate using the Molecular Complex Detection (MCODE) algorithm. MCODE analysis was carried out using the Cytoscape program by k-core (2), degree cutoff (2), a maximum depth (100), node score cutoff (0.2). These helped us identify key biological pathways and subnetworks relevant to HCC progression.

### Establishment and validation of prognostic risk model

LASSO regression analysis was conducted on the 75 node genes screened out by the MCODE algorithm, and selected the minimum lambda value (lambda. min=0.0166) in this study. To create a prognostic risk model, a risk score was determined for each TCGA-HCC tumor sample. We created scatterplots and heatmaps of gene expression for the risk model and identified 26 significant genes. The TCGA samples were split into high-(n=185) and low-risk (n=185) groups according to average risk score, and the overall survival (OS) analyses was performed by Kaplan-Meier database. The risk score formula was as follows: Riskscore = (-0.0199)*RPA1 + (0.2099)*WDR12 + (0.0206)*CDK4 + (0.0147)*DCAF13 + (0.1308)*FTSJ3+(0.3767)*CPSF3 + (-0.1757)*RFC1 + (-0.1788)*RRP1B + (0.1366)*RMI1 + (-0.0077)*POLD3 + (0.1838)*ASF1A + (0.1581)*CPSF2 + (0.0078)*NUP43 + (0.1364)*XPOT + (0.0332)*CCNF + (0.1683)*ESF1 + (0.23IP1) + (-0.0111)*NOL12 + (-0.1896)*DDX59 + (-0.4887)*RFC3 + (0.487)*TTK + (0.0187)*RRM2 + (0.0052)*CEP55 + (0.0855)*ECT2 + (0.0763)*FBXO5 + (-0.6687)*ZWILCH. Then, using receiver operating characteristic (ROC) analysis, we reported 1-, 3-, and 5-year OS and compared OS curves between subgroups. The capability of the model was evaluated by area under the area under curve (AUC).

### Prognostic gene expression and survival analysis in TCGA and GEO

First, we obtained gene data from TCGA, GSE45267 and GSE49515 datasets, respectively. Then, univariate Cox regression analysis was employed to determine the most important prognostic genes. Next, we applied batch survival analysis on the selected genes to identify genes significantly related to patient survival, by Kaplan-Meier tool.

### Prognostic nomogram construction

The Cox proportional hazards model is a widely applied statistical method for survival analysis, which considers the influence of multiple variables on the survival time of patients. Herein, we applied univariate/multivariate Cox analyzes to investigate the prognostic value of 19 genes with significant expression in survival analysis. Multivariate Cox analysis was used to assess the individual prognostic potential of each gene after adjusting for other covariates. Finally, we selected genes substantially related to survival outcomes in both univariate and multivariate Cox analyzes to construct prognostic nomograms. The nomogram was further validated using a calibration curve.

### Clinical feature analysis of key gene and immune score evaluation

In this study, we utilized the UALCAN (http://ualcan.path.uab.edu/index.html) database to study the levels and trends of key genes in the clinical characteristics of liver cancer, including age, gender, individual cancer, TP53 mutation and tumor grade. UALCAN is an interactive portal that provides easy access to TCGA data. The immune scores were then further evaluated using immuneDeconv that uses gene expression data to evaluate the comparative abundance of immune cell types in tumor samples.

### Cell culture

American type culture collection (ATCC) provided HCC cell lines (Huh7, Hep3B, HepG2, MHCC97H) and normal liver cell (LO2). HepG2 and MHCC97H were put in RPMI-1640 media with 10% FBS, whereas Huh7 and Hep3B in DMEM/F12 conditions. 10% FBS was added to DMEM to boost the culture of LO2 cells. Experiments were conducted on cells between passages 3 and 8, and cells were subcultured every three to four days.

### Quantitative reverse transcription polymerase chain reaction (qRT-PCR)

We conducted siRNA transfection by Lipofectamine RNAiMAX Transfection Reagent. Total RNA was collected from the cells 48 hours after transfection by TRIzol reagent. Utilizing the StepOnePlus Real-Time PCR System and SYBR Green PCR Master Mix, qRT-PCR analysis was carried out. The primers for PAK1IP1 were as follows: forward 5’-TGGTCCACGATGCCCTATG-3’ and reverse 5’-GGCTTTGGT TTCGGTGTTGT-3’; for GAPDH: forward 5’-ACAGTCAGCCGCATCTTCTT-3’ and reverse 5’-GTTAAAAGCAGCCCTGGTGA-3’.

### Western blotting (WB)

Using RIPA buffer enhanced with protease and phosphatase inhibitors, total protein was recovered from cells. The BCA protein assay kit was applied to calculate the protein concentration. Electrophoresis was used to separate equal quantities of protein (20-40 µg) that had been put onto 10-12% SDS-PAGE gels. A wet transfer technique was then employed to transfer the isolated proteins onto nitrocellulose or PVDF membranes. Membranes were kept with primary antibodies at 4°C overnight after blocked with 3% BSA in TBST for an hour. The membranes were TBST-washed before incubation for an hour with secondary antibodies that were HRP-conjugated. An ECL substrate was used to see the protein bands, and an imaging equipment was used to take pictures of them. For the purpose of balancing the amounts of protein expression, we employed either β-actin or GAPDH as an internal control. With ImageJ, the bands were quantified.

### Cell counting kit-8 (CCK-8) assay

5×10^3^ cells were planted each well in 96-well plates, and the cells were then left to attach for 24 hours. After treatment with the experimental reagents, the CCK-8 solution was diluted to 10 L in each well, and the plates were incubated at 37°C for 1-4 hours. The absorbance at 450 nm was measured with a microplate reader. By contrasting the absorbance of treated and control cells, the vitality of the cells was determined. The experiments were carried out three times, and the mean standard deviation was used to show the findings.

### Transwell assay

Cells were put into the upper chamber of a Transwell insert with an 8 µm hole size for the migration test after being suspended in serum-free DMEM. 10% FBS-containing media was put into the bottom chamber. The upper chamber of the Transwell insert had Matrigel (BD Biosciences) precoated for use in the invasion experiment. In order to seed the upper chamber, cells were suspended in serum-free media. 10% FBS-containing media was put into the bottom chamber. The migrating and invading cells were fixed and stained with DAPI. The amount of invading or migrating cells was counted after microscopically captured images.

### ELISA assay

To assess the amounts of IL-1 protein expression in cell culture supernatants, the enzyme-linked immunosorbent assay (ELISA) was used. Briefly at 4°C, 96-well plates were coated with anti-IL-1β capture antibody overnight. The wells were filled with the cell culture supernatants and cultured for 2 hours indoor after blocking with 1% bovine serum albumin in Phosphate buffered saline (PBS) for 2 hours. Anti-IL-1 detection antibody that had been biotinylated was then added to the wells and incubated for an additional hour indoor in a solution containing 1% bovine serum albumin and PBS. Streptavidin-horseradish peroxidase (1:5000 dilution) was added to each well after three PBS washes with 0.05% Tween 20 were completed. Each well was then kept for 30 minutes. Finally, the wells were rinsed by PBS with 0.05% Tween 20 and the reaction was developed using tetramethylbenzidine substrate solution. The optical density was detected at 450 nm by microplate reader. The content of IL-1β in the cell culture supernatants was computed based on the standard curve. All samples were run in duplicate.

### Flow cytometry analysis

Cells were collected and quickly cleaned with cold PBS. Cells were frozen with 70% ethanol at -20°C for 24 hours, stained with propidium iodide (PI), and left in the dark for 30 minutes to analyze the cell cycle. For apoptosis analysis, the manufacturer’s recommendations were followed while staining by Annexin V-FITC/PI apoptosis detection kit. Then, cells were investigated by flow cytometer (BD FACS Calibur). The proportion of cells that underwent apoptosis in each group was used to display the results. After data collection, we performed detailed data analysis and graph generation using FlowJo software (version 10.6.1, FlowJo, LLC, USA).

### Statistical analysis

Various bioinformatics and statistical analysis packages of R language were employed, such as edgeR, limma, survival, ggplot2, etc. We preprocessed each dataset, including normalization of gene expression levels and correction for batch effects. For the screening of DEGs, we applied the edgeR and limma packages to screen for differentially expressed genes, and set the significance level as FDR<0.05 and log_2_ fold change >1 or <-1.

## Results

### Identification of DEGs and key module in WGCNA

We performed a comprehensive analysis of liver cancer using a multi-database integration. Totally, we found 8977 up- and 1001 down-regulated DEGs from the TCGA dataset (Figure 1A), 4717 up- and 2343 down-regulated DEGs from the GSE45267 dataset (Figure 1B), and 3442 up and 1682 down-regulated DEGs (Figure 1C). The Venn diagram revealed 1163 upregulated genes and 76 downregulated genes at the intersection (Figures 1D and 1E). By using WGCNA analysis, we determined a soft threshold power of 1 (Figure 1F). Two modules were identified based on the clustering dendrogram (Figure 1G), and measured the relation between module eigengenes (ME) and clinical characteristics, the turquoise module was the key module for subsequent analysis (Figure 1H).

**Figure 1.**
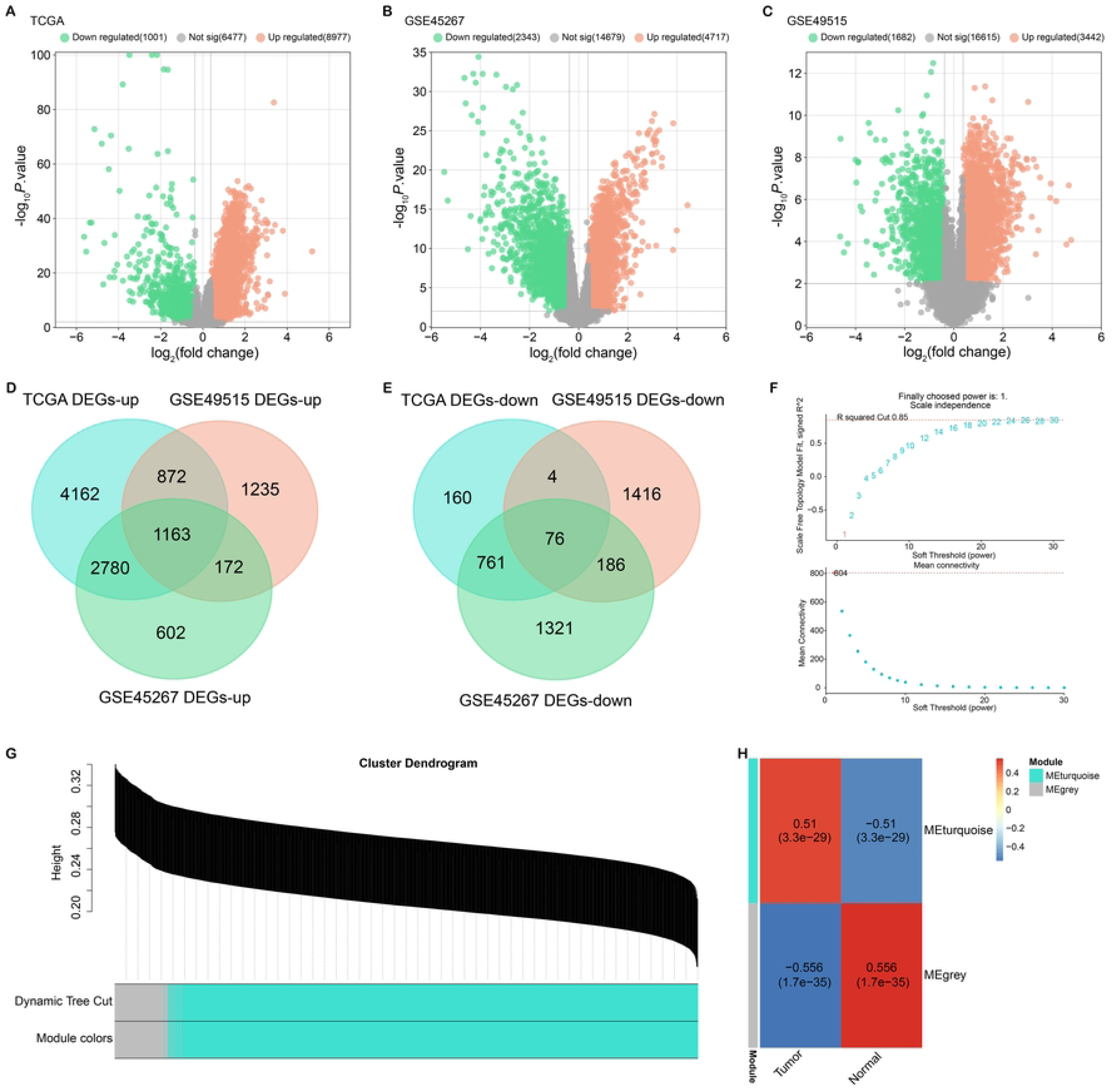
Identification and WGCNA analysis of DEGs in liver cancer. (A-C) Volcano plots showing identified DEGs from the TCGA dataset (A), GSE45267 dataset (B) and GSE36376 dataset (C). Orange and green dots represent up-regulated and down-regulated genes, respectively. (D) Venn diagram showing the intersection of upregulated DEGs from the three datasets. (E) Venn diagram showing the intersection of downregulated DEGs from the three datasets. (F) Determination of soft threshold power using WGCNA analysis. (G) Dendrogram of gene clustering in WGCNA analysis, with modules indicated by color. (H) Correlation heatmap showing correlations between modular eigengenes (MEs) and clinical features. The Turquoise module was identified as a key module related to clinical features.

### The functional enrichment analysis and PPI networks of the turquoise module

The genes in turquoise module were related to DNA metabolic process, Mitotic spindle organization, and DNA repair (Figure 2A). Furthermore, enriched pathways included RNA transport, Herpes simplex virus 1 infection, Cell cycle, and DNA replication (Figure 2B). Additionally, we utilized the MCODE algorithm to analyze the PPI network of the turquoise module genes. Figures 2C and 2D, respectively, display the mcode1 cluster (30 nodes and 379 edges) and the mcode2 cluster (45 nodes and 247 edges). In summary, our findings suggested that the turquoise module genes are associated with crucial biological processes and pathways involved in HCC tumorigenesis.

**Figure 2.**
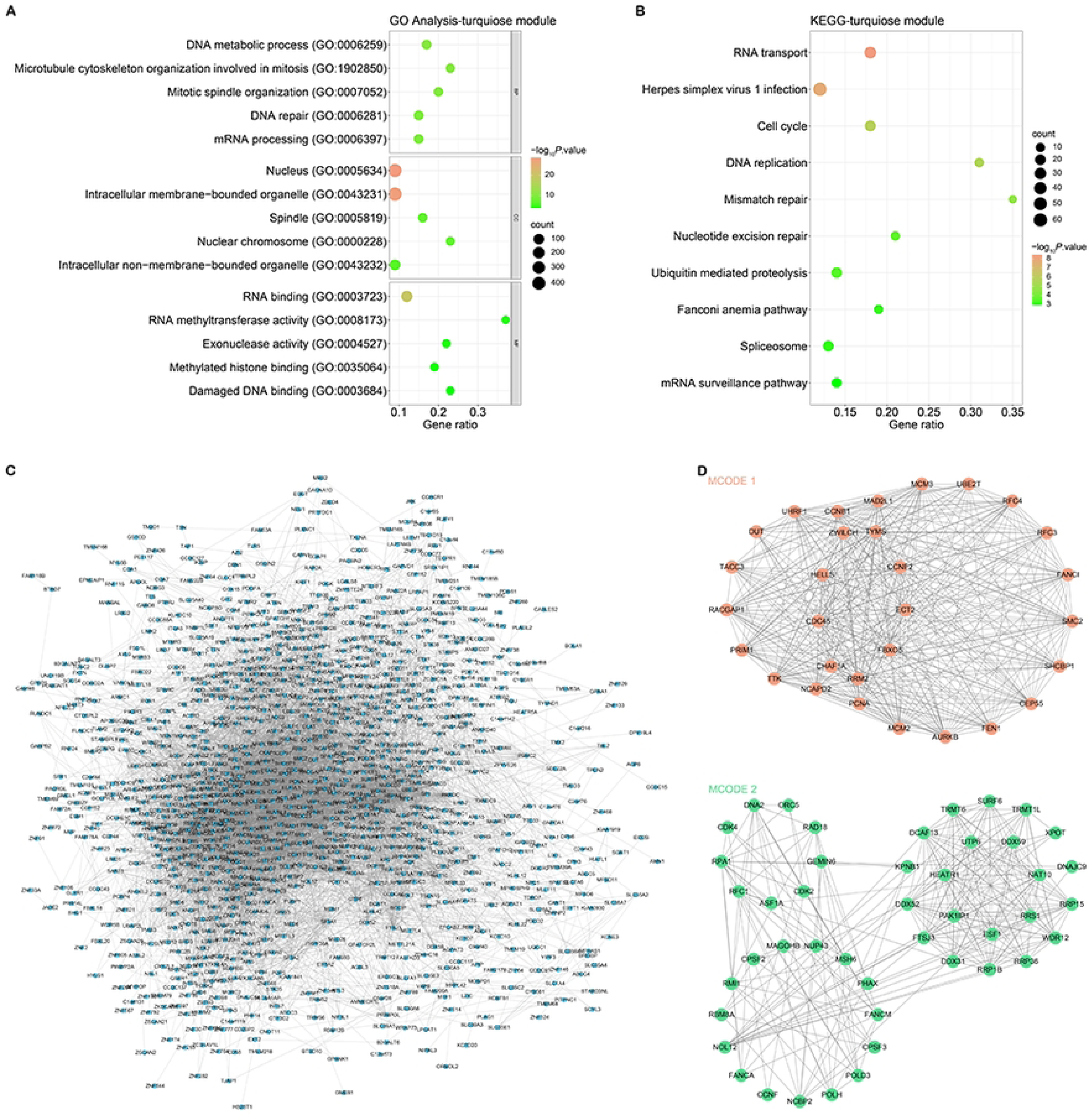
GO, KEGG pathway enrichment analysis and MCODE networks of turquoise module genes. (A) GO analysis of turquoise module gene enrichment. The abscissa is the Gene ratio, and the ordinate is the enriched term. (B) KEGG pathway enrichment analysis of turquoise module genes. The abscissa is the Gene ratio, and the ordinate is the enriched pathway. (C) PPI network of the mcode1 cluster within the turquoise module gene. (D) PPI network of the mcode2 cluster within the turquoise module gene. Nodes in the network represent genes and edges represent protein-protein interactions. The size of the nodes reflects the degree of connectivity of the corresponding genes.

### Identification of prognostic genes through construction of risk model for HCC

In this study, we conducted LASSO regression analysis on nodal genes and determined an optimal lambda.min value of 0.0166 (Figures 3A and 3B). Based on this value, we developed a risk model for liver cancer and identified 26 prognostic genes that exhibited statistical significance. The 26 gene level in HCC samples was shown in Figure 3C, with higher risk scores linking to higher mortality. In addition, the OS analysis revealed the high-risk group’s survival prognosis was poorer (Figure 3D). Besides, ROC analysis showed AUC values of 0.858, 0.79, and 0.749 for the first, third, and fifth years, respectively (Figure 3E). These findings provided valuable insights into HCC prognosis and highlighted the potential clinical utility of the identified prognostic genes and risk models.

**Figure 3.**
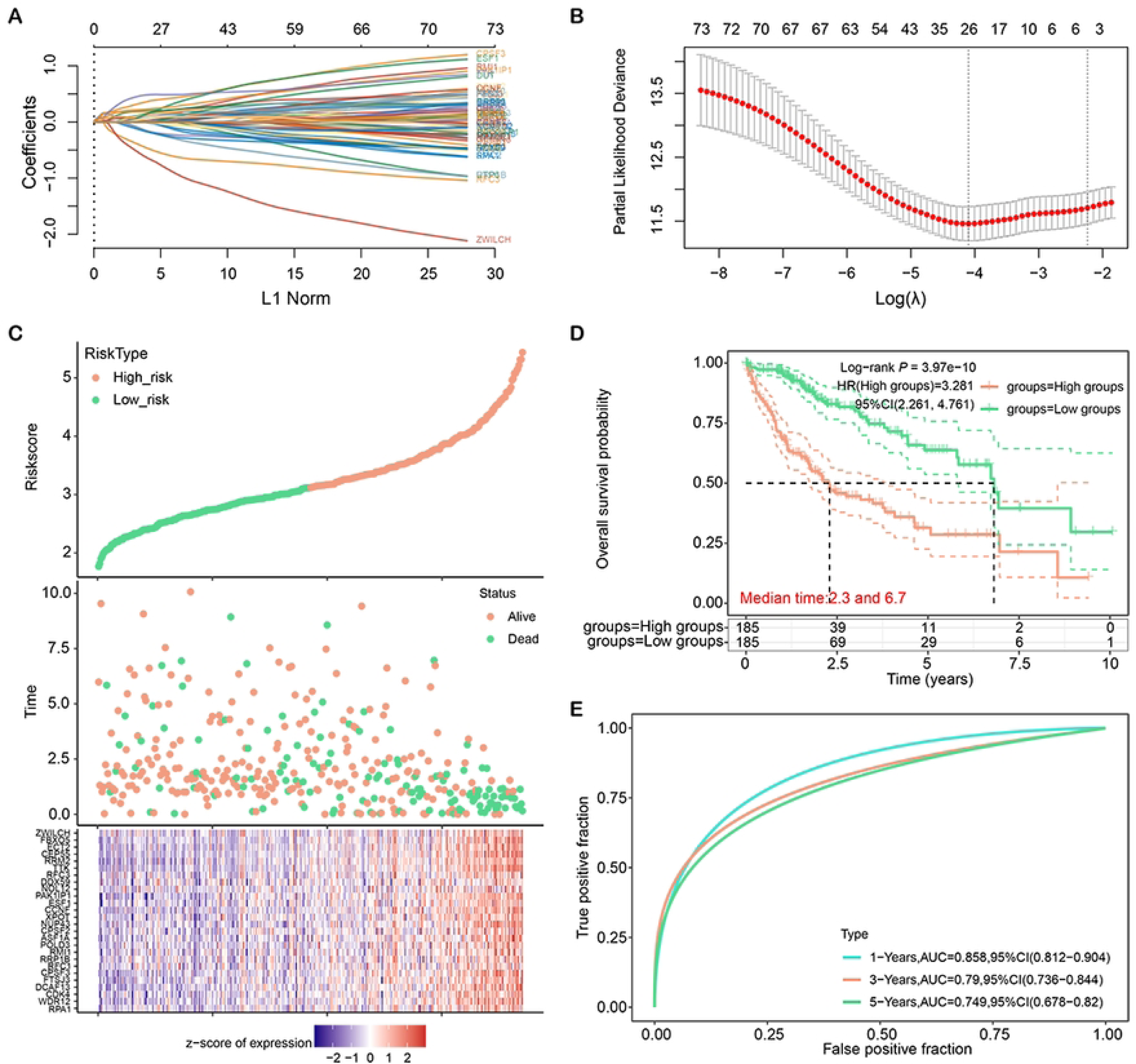
LASSO regression analysis and construction of liver cancer risk model. (A) Distribution plot of LASSO coefficients for nodal genes. (B) Optimal lambda.min value selected by 10-fold cross-validation. (C) Patient characteristics ordered by their risk score. From top to bottom are the risk scores of 26 genes, the distribution of patient survival status, and the heat map of patients in the low-risk group and high-risk group. (D) Kaplan-Meier curves of overall survival in high-risk and low-risk groups. (E) ROC curves of the liver cancer risk model at 1, 3, and 5 years.

### 19 prognostic genes associated with survival in HCC patients

We investigated the expressions of 26 prognostic genes in TCGA, GSE45267, and GSE49515 datasets (Figures 4A-4C). Our results demonstrated these genes were upregulated in tumor samples. Kaplan-Meier survival analysis indicated 19 genes were significantly linked to poor prognosis, and high expressions demonstrated lower survival (Figures 4D-4V). Taken together, these 19 genes have potential as prognostic biomarkers for cancer patients and can be used to develop personalized treatment strategies.

**Figure 4.**
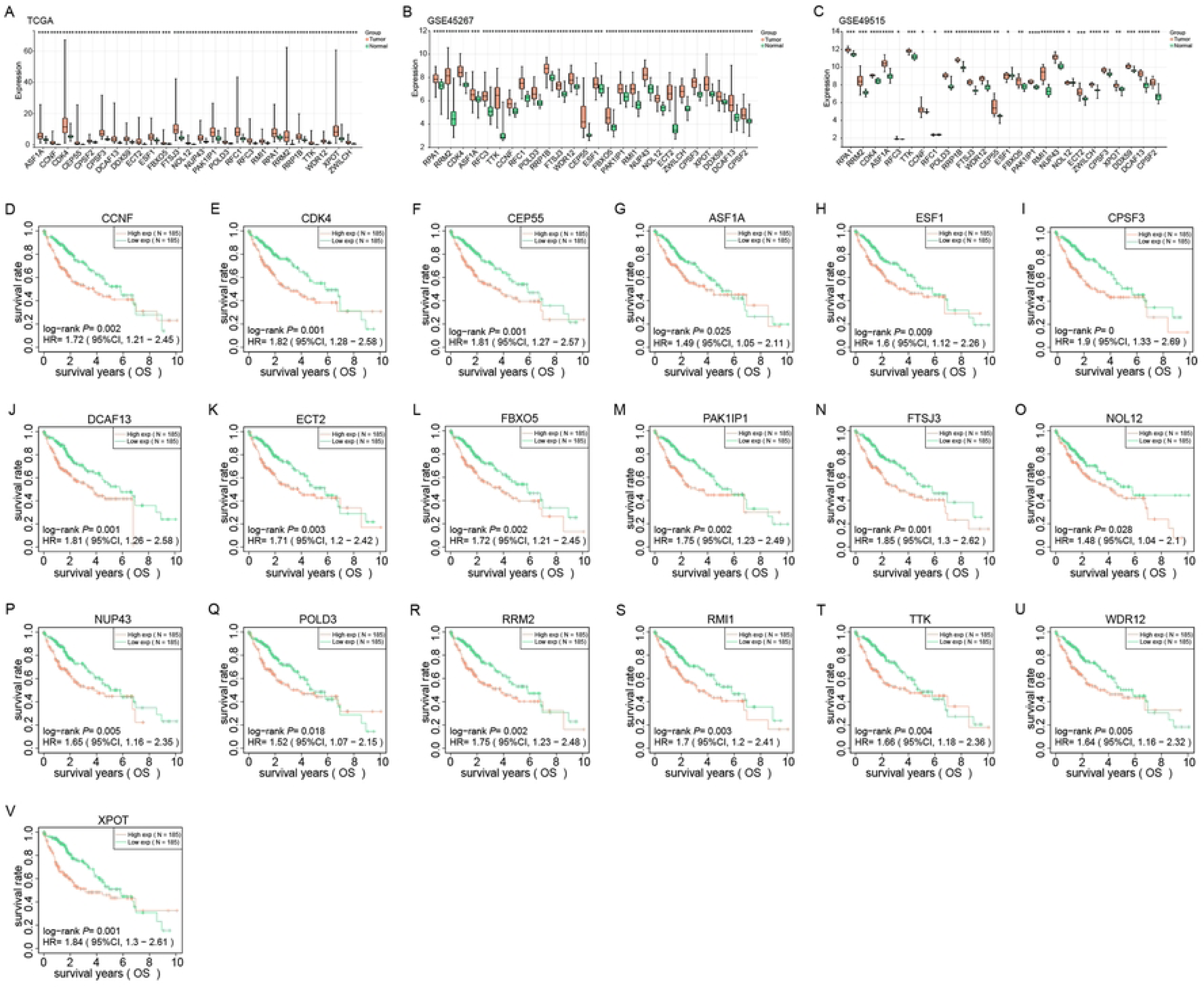
Expression and prognostic analysis of 26 identified genes in liver cancer. (A-C) Expression levels of 26 genes from TCGA, GSE45267 and GSE49515 datasets in normal and tumor tissues. Red and green represent tumor and normal samples, respectively. (D-V) Kaplan-Meier survival analysis of 26 genes in HCC patients in the TCGA dataset. The X-axis represents the survival time, and the Y-axis represents the survival rate. Red and green curves represent high and low expression groups, respectively. *P*-values and hazard ratios (HR) with 95% confidence intervals (CI) are shown. \**P*<0.05, \*\**P*<0.01, \*\*\**P*<0.001, \*\*\*\**P*<0.0001.

### PAK1IP1 is an individual prognostic gene for HCC patients

In this study, we used Cox regression analysis, both univariate and multivariate, and discovered that PAK1IP1 and pTNM stage were individual predictive variables for HCC (Figures 5A-5B). We then integrated PAK1IP1 and pTNM stage to build a nomogram for more accurate prognosis prediction in HCC patients (Figure 5C). The calibration plot revealed that the projected and actual survival rates were well-aligned (Figure 5D). These emphasized the value of these two factors in HCC prognosis and the potential of integrating them in a nomogram for clinical use.

**Figure 5.**
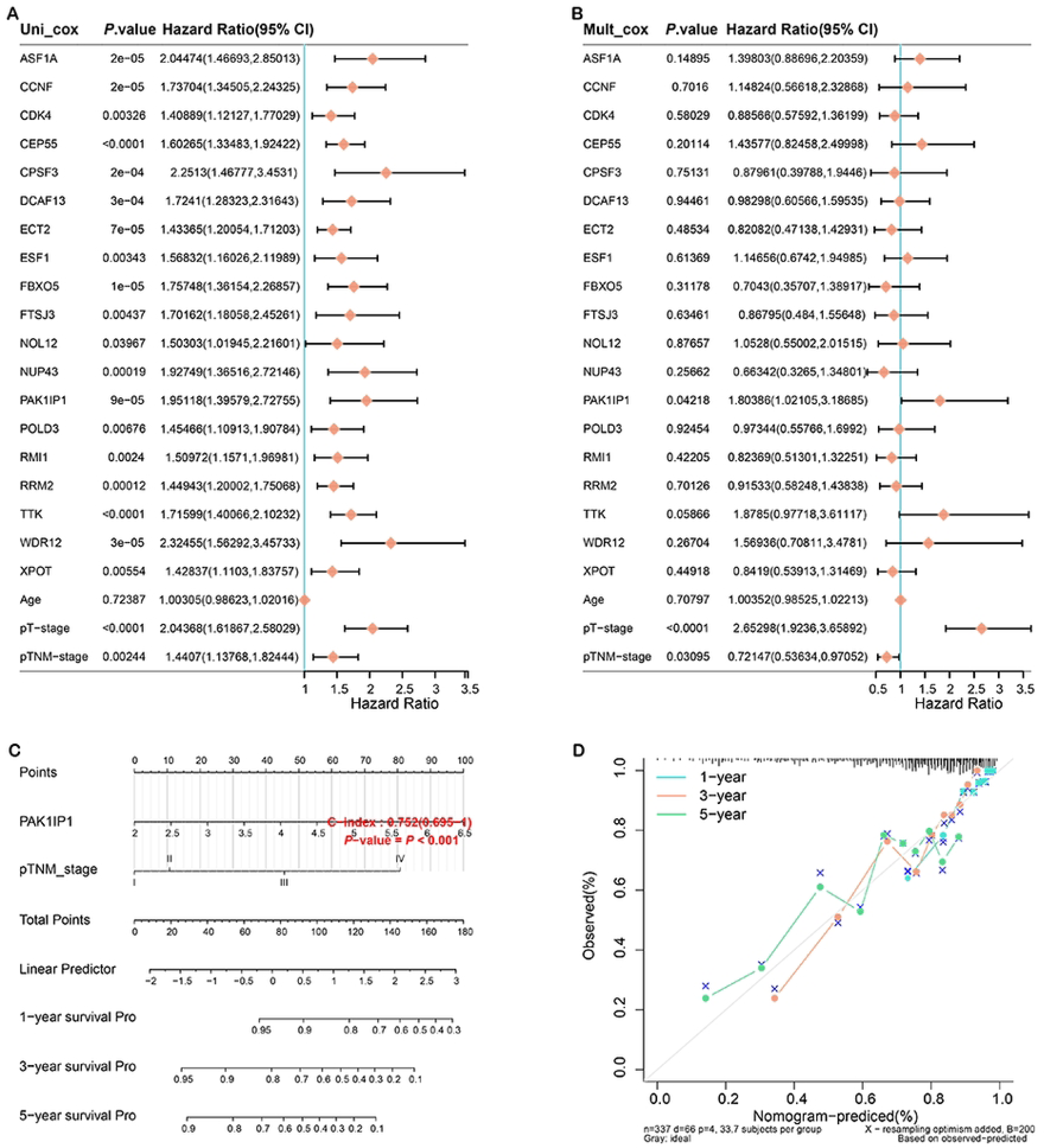
Construction of the prognostic nomogram. (A) Forest plot of univariate Cox regression analysis of PAK1IP1 and clinicopathological variables. (B) Forest plot of multivariate Cox regression analysis of PAK1IP1 and clinicopathological variables. (C) Nomogram for HCC prognosis prediction constructed by integrating PAK1IP1 and pTNM staging. (D) Calibration plot of the nomogram showing the agreement between predicted and observed survival. The x-axis represents predicted survival probabilities and the y-axis represents actual survival probabilities. Diagonal lines represent perfect predictions.

### PAK1IP1 expression correlates with immune cell infiltration in HCC

Next, we studied the level and clinical significance of PAK1IP1 in LIHC. According to data from the UALCAN database, PAK1IP1 level was considerably greater in tumor samples (Figure 6A). Furthermore, PAK1IP1 level was higher in HCC patients with older age, male gender, and TP53 mutation (Figures 6B, C, E). In addition, as tumor stage and grade increased, PAK1IP1 expression also increased (Figures 6D, F). Next, we identified most immune cells were downregulated in the low PAK1IP1 expression group, and Myeloid dendritic cells had the highest infiltration percentage in tumor samples (Figures 6G-6H). Our findings suggested that PAK1IP1 may be a predictive biomarker and be connected to immune cell infiltration.

**Figure 6.**
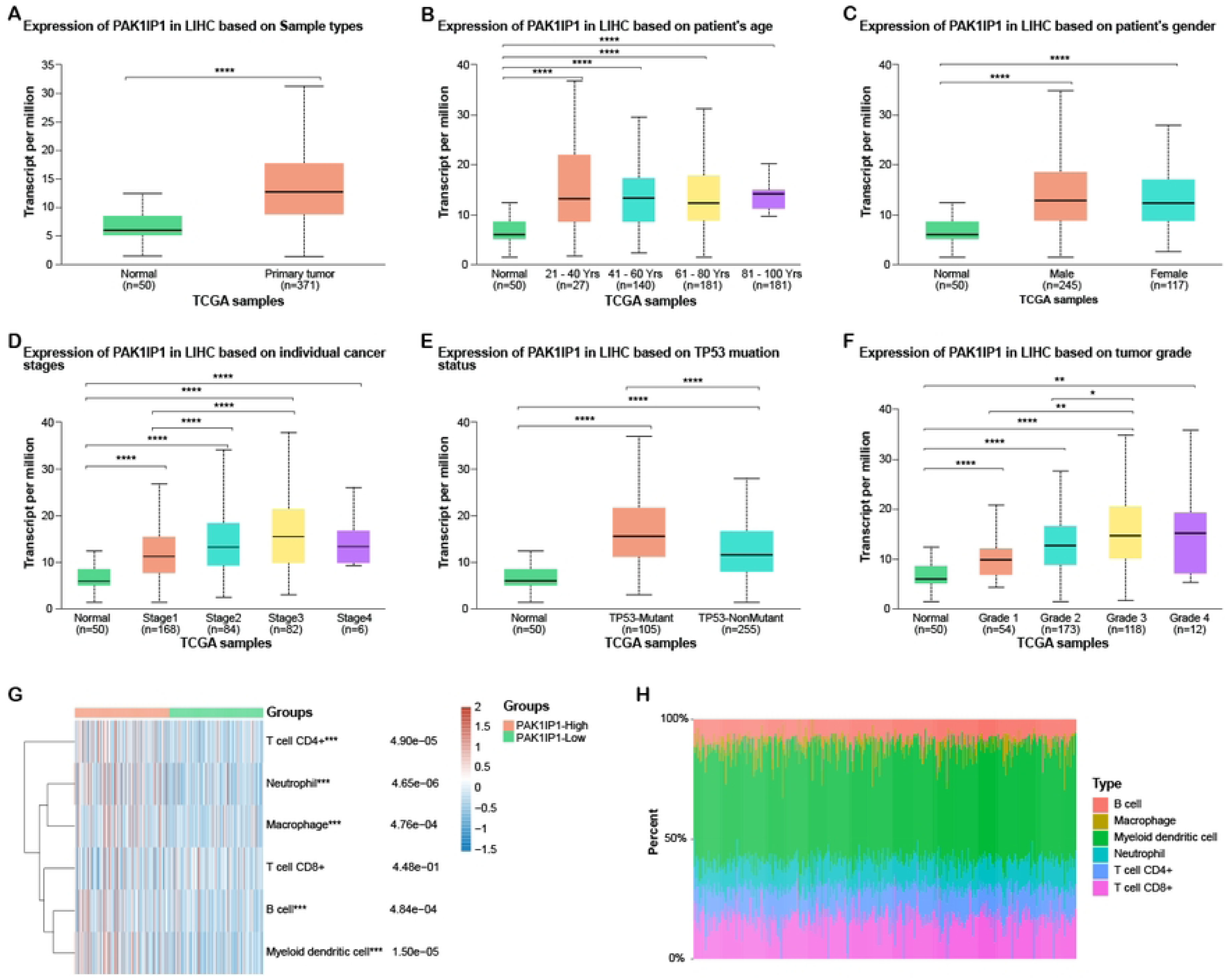
Clinical feature and immune analysis of PAK1IP1 in liver cancer. (A) PAK1IP1 expression levels in normal and tumor samples according to the UALCAN database. (B-F) Boxplot displaying PAK1IP1 expression levels in individuals with liver cancer, stratified by (B) age, (C) sex, (D) tumor stage, (E) TP53 mutation status, and (F) tumor grade. (G) Heat map of immune cell scores, where different colors represent expression trends in different samples. (H) The percentage abundance of tumor-infiltrating immune cells in each sample. Different colors represent different immune cell types, the abscissa represents the sample, and the ordinate represents the percentage of immune cell content in a single sample. \**P*<0.05, \*\**P*<0.01, \*\*\*\**P*<0.0001.

### PAK1IP1 is up-regulated in HCC cells

We discovered through qRT-PCR analysis that HCC cells (especially Hep3B and HepG2) have higher levels of PAK1IP1 expression than normal liver cells (Figure 7A). Our results demonstrated that si-PAK1IP1#1 and si-PAK1IP1#2 were more efficient in reducing PAK1IP1 expression in HCC cells (Figure 7B). Moreover, protein analysis by western blotting confirmed that si-PAK1IP1#1 and si-PAK1IP1#2 significantly reduced PAK1IP1 expression in HCC cells (Figure 7C). We then applied CCK-8 assays to examine the influence of PAK1IP1 knockdown on HCC cell growth. As expected, PAK1IP1 knockdown inhibited HCC cell growth (Figures 7D-7E). Additionally, Transwell assays showed that PAK1IP1 knockdown significantly suppressed HCC cell invasion and migration (Figures 7F-7I).

**Figure 7.**
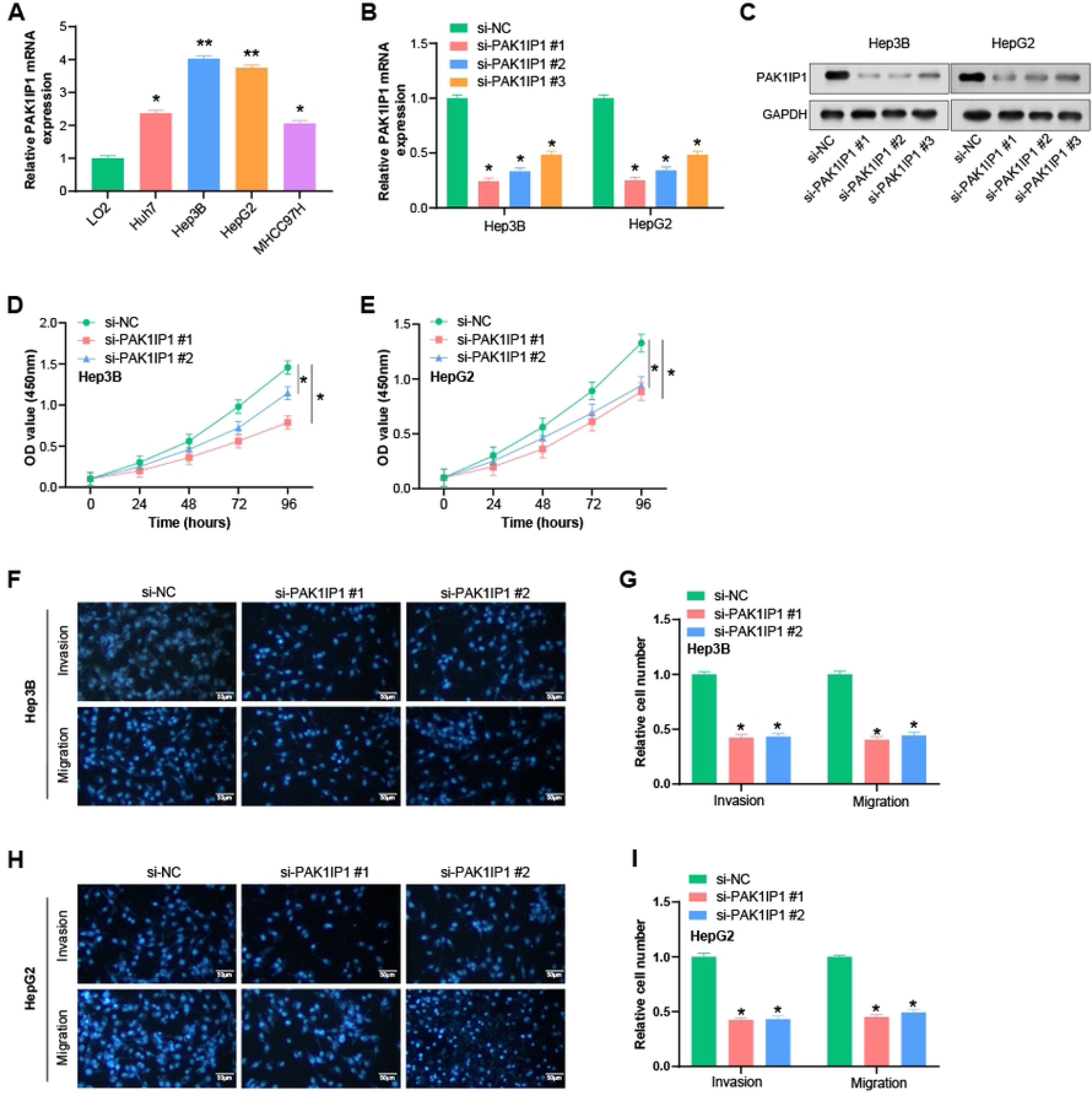
Knockdown of PAK1IP1 inhibits growth, invasion and migration of HCC cells. (A) qRT-PCR analysis of PAK1IP1 expression in normal hepatocytes and HCC cells. (B-C) Efficiency of si-PAK1IP1 #1, #2 and #3 in HCC cells detected by PCR and WB in HCC cells. GAPDH was used as a loading control. (D-E) CCK-8 assay showed the effect of PAK1IP1 knockdown on the proliferation of HCC cells. (F-I) Transwell assay for the effect of PAK1IP1 knockdown on the invasion and migration of HCC cells. Scale: 50 µm. \**P*<0.05, \*\**P*<0.01.

### PAK1IP1 knockdown could activate lipopolysaccharide (LPS)-induced pyroptosis

We constructed a physical interaction network between PAK1IP1 and 33 pyroptosis-related genes from previous research[15] (Figure 8A). To investigate whether PAK1IP1 was related to the regulation of pyroptosis, we measured the production of IL-1β, a key marker of pyroptosis, in HCC cells after LPS treatment and PAK1IP1 knockdown using ELISA. According to our findings, IL-1 levels considerably increased following LPS treatment and PAK1IP1 knockdown (Figures 8B-8C). Flow cytometry also demonstrated an up-regulation in the apoptosis rate of HCC cells following LPS treatment and PAK1IP1 knockdown (Figures 8D-8E).

**Figure 8.**
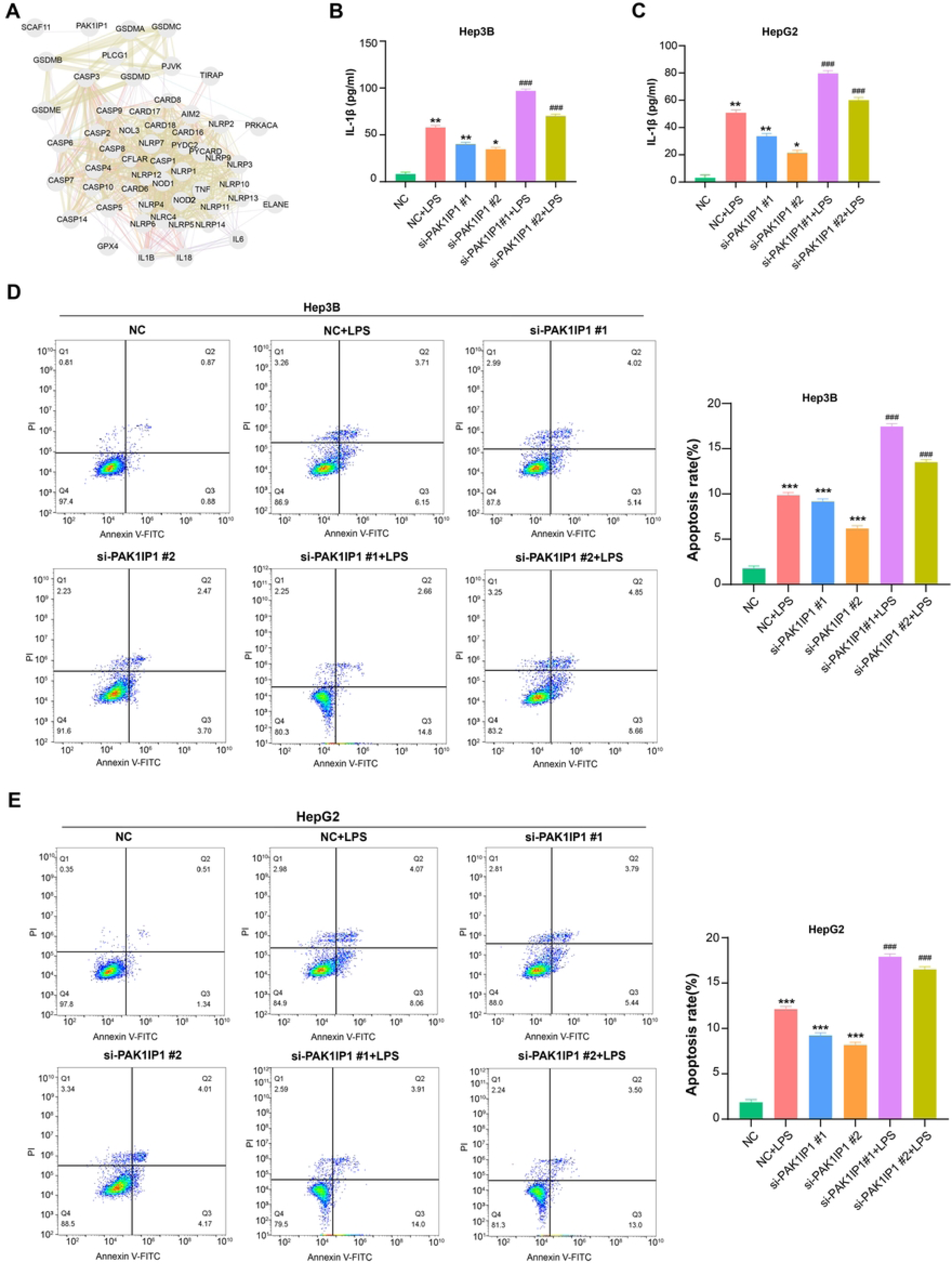
PAK1IP1 regulates gene expression associated with pyroptosis in HCC cells. (A) Physical interaction network between PAK1IP1 and 33 pyroptosis-related genes. Nodes represent genes, and edges represent connectivity between genes. (B-C) ELISA assay to detect the effect of LPS treatment and knockdown of PAK1IP1 hepatocellular carcinoma cells on pyroptosis-related markers (IL-1β). (D-E) The effect of LPS treatment and PAK1IP1 knockdown on the apoptosis rate of HCC cells was determined by flow cytometry. Quantitative results were shown on the right side of the graph. \**P*<0.05, \*\**P*<0.01, \*\*\**P*<0.001.

### PAK1IP1 promotes HCC proliferation, invasion and migration through suppressing the CASP-3-dependent pyroptosis

Moreover, we found that PAK1IP1 inhibition significantly increased the protein levels of CASP-3, as detected by WB (Figure 9A). We then assessed the effects of si-PAK1IP1 #1 and Z-DEVD-FMK treatment, a CASP-3 inhibitor, on liver cancer cell proliferation. The data showed that PAK1IP1 knockdown significantly reduced HCC cell proliferation, while Z-DEVD-FMK treatment increased cell proliferation (Figures 9B-9C). Transwell assays demonstrated that PAK1IP1 knockdown suppressed liver cancer cell invasion and migration after LPS treatment, while Z-DEVD-FMK treatment promoted cell invasion and migration (Figures 9D-9G). These findings indicate PAK1IP1 is a vital part in HCC cell proliferation, invasion, and migration through the CASP-3 pathway.

**Figure 9.**
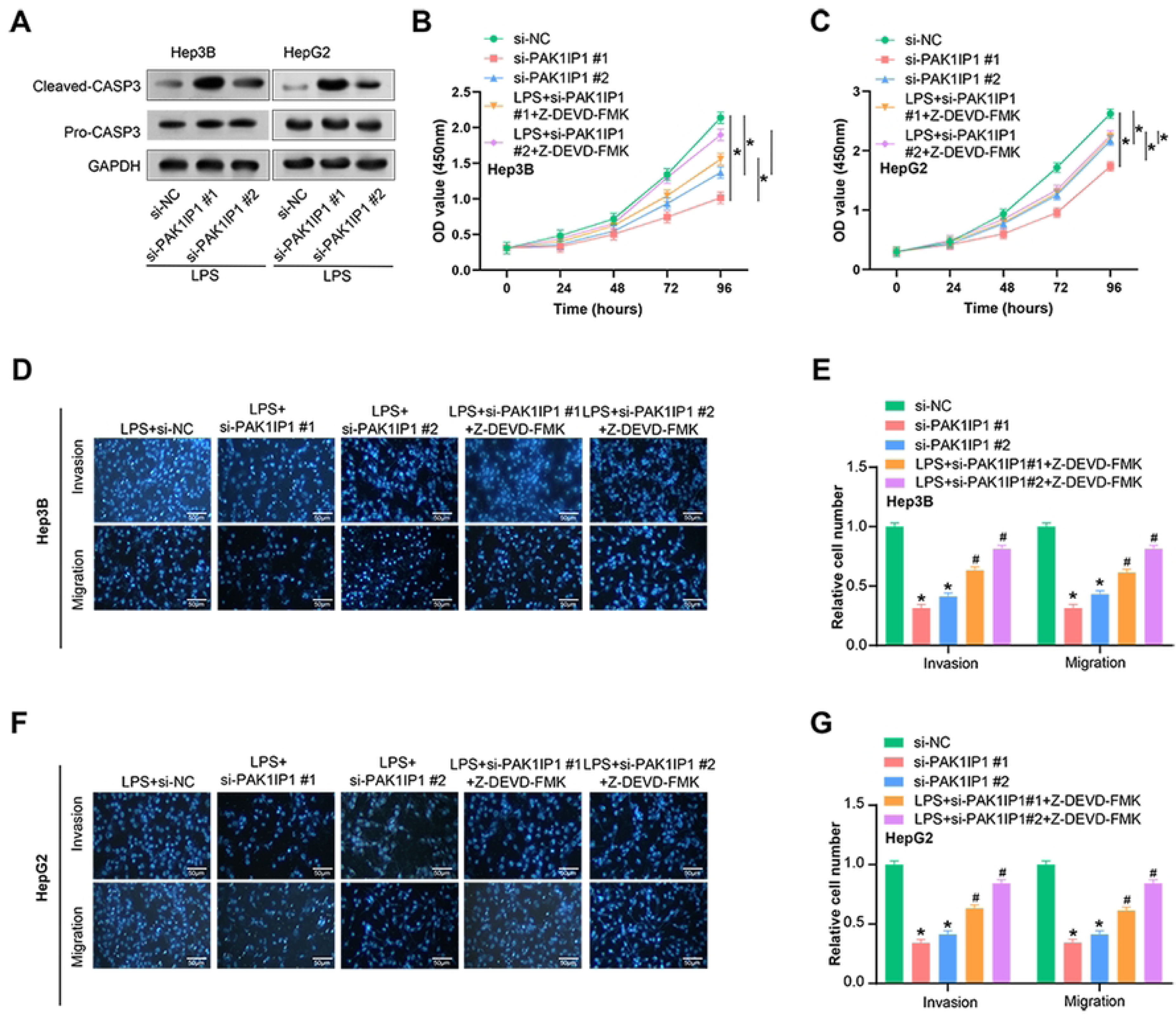
PAK1IP1 knockdown suppresses proliferation, invasion and migration of HCC cells through the CASP-3 pathway. (A) Western blot analysis of CASP-3 protein expression in HCC cells after PAK1IP1 knockdown. (B-C) CCK-8 assay showing the effect of si-PAK1IP1 knockdown and Z-DEVD-FMK treatment on the proliferation of HCC cells. (D-E) Transwell assay showing the effect of PAK1IP1 knockdown on the invasion and migration of HCC cells after LPS treatment. (F-G) Transwell analysis showing the effect of Z-DEVD-FMK treatment on the invasion and migration of HCC cells. Scale: 50 µm. \**P*<0.05.

## Discussion

Biomarkers are a crucial part in the diagnosis, therapy, and prognosis of cancer[16]. Currently, the diagnosis of liver cancer heavily relies on CT, MRI and others[17, 18]. However, these methods have limitations in detecting early-stage tumors. To address this challenge, targeted gene therapy has become a new therapy for HCC[19]. Nevertheless, its efficiency is hindered by tumor heterogeneity and the development of acquired resistance[20]. Furthermore, the five-year survival rate for liver cancer remains low because of high recurrence and metastasis. Consequently, exploring new efficient biomarkers for the detection, management, and prognosis of liver cancer is urgently needed. These biomarkers could facilitate early detection, guide personalized treatment strategies, and predict patient outcomes. Ultimately, the identification of such biomarkers would significantly improve the survival of HCC patients.

In our study, we identified the turquoise module as an important module associated with liver cancer. The genes in the turquoise module were primarily related to DNA metabolic processes, mRNA processing, nuclear chromosome function, RNA methyltransferase activity, methylated histone binding, Fanconi Anemia Pathway, and mRNA Surveillance Pathway. These enriched pathways and terms have been extensively studied in HCC and have shown promising results in improving our understanding of the disease. One of the pathways enriched in the turquoise module is the DNA metabolic process, which plays a crucial role in DNA replication, recombination, and repair[21]. Dysregulation of this pathway has been linked to various cancers, including liver cancer[22]. For example, a study found that the expression of the DNA polymerase kappa (POLK) gene involved in DNA metabolism was upregulated in liver cancer tissues[23]. Another enriched pathway in the turquoise module is the cell cycle. A study reported that when compared to nearby normal tissues, cell cycle pathway gene CDKN2A was considerably downregulated in liver cancer tissues[24]. Furthermore, low CDKN2A level is linked with a poor prognosis in HCC patients[25]. The mismatch repair pathway, involved in correcting errors during DNA replication, is also enriched in the turquoise module. Defects in this pathway have been related to the progression of various cancers. For instance, a study reported that the expression of the MutS homolog 2 (MSH2) gene involved in the mismatch repair pathway was downregulated in liver cancer tissues[26]. Additionally, patients with HCC who have low MSH2 expression have a bad prognosis[27]. Overall, the enriched pathways and terms in the HCC turquoise module give insightful information on the molecular pathways underlying the genesis and progression of diseases. Further research on these pathways may help identify potential targets for new therapies in HCC.

Based on the above results and a series of bioinformatics analysis, the key gene PAK1IP1 was determined in this study. PAK1IP1 exerted crucial regulatory functions in various biological processes[28]. Aberrant expression of PAK1IP1 has been implicated in human diseases[29]. PAK1IP1 interacts with β-catenin and facilitates its nuclear translocation, resulting in the activation of Wnt target genes[30]. Similarly, in gastric cancer, PAK1IP1 enhances cell invasiveness by regulating actin cytoskeleton dynamics[31]. Furthermore, PAK1IP1 has been implicated in cancer cell survival and chemotherapy resistance[32]. In ovarian cancer, PAK1IP1 promotes cell survival under hypoxic conditions by regulating the AMPK signaling pathway. PAK1IP1 binds to and inhibits the activity of AMPK, thereby reducing cell death and promoting cell survival[28]. In our research, we found that HCC tumor tissue samples had a high expression of PAK1IP1. Moreover, clinical tissue samples from liver cancer patients exhibited upregulated expression of PAK1IP1. The PAK1IP1 high-expression group had a poorer OS rate, according to the Kaplan-Meier survival analysis, compared to the low-expression group. Overall, PAK1IP1 acts as a proto-oncogene in liver cancer patients, and its overexpression adversely affects patient outcomes. Understanding the molecular processes that underlie PAK1IP1-mediated cancer growth may offer fresh perspectives for the creation of fresh treatment approaches.

Myeloid dendritic cells (mDCs) have been identified as crucial players in the immune response against cancer[33]. Herein, we found a significant increase in mDC infiltration in HCC patients. These mDCs function as antigen-presenting cells, initiating the activation of T cells to recognize and eliminate cancer cells[34]. Additionally, they contribute to the activation of natural killer (NK) cells, which are essential for tumor surveillance[35]. Moreover, mDCs can generate cytokines that facilitate the recruitment and activation of other immune cells like macrophages and neutrophils, further bolstering the antitumor immune response[36]. In liver cancer, mDCs have been found to be a critical part in initiating and enhancing antitumor immune responses[37]. Notably, mDCs promote the differentiation and activation of CD8+ T cells, which are instrumental in eliminating tumor cells[38]. Furthermore, mDCs produce interleukin 12 (IL-12), which augments the antitumor activity of NK cells[39]. However, it should be noted that the role of mDCs in tumor growth promotion and immune evasion has also been reported in certain types of cancer, necessitating further investigations to fully comprehend their intricate and multifaceted functions in cancer biology. In conclusion, our study underscores the potential significance of mDCs in liver cancer and indicates targeting mDCs could be a promising method to enhance antitumor immune responses. Subsequent studies are imperative to unravel the mechanisms underlying mDC function in cancer and to develop novel immunotherapeutic approaches aimed at harnessing the potential of these cells.

Furthermore, we conducted experiments to elucidate the mechanism of PAK1IP1 and pyroptosis-related genes in liver cancer. To assess pyroptosis, we utilized ELISA to measure the production of IL-1β, a key pyroptosis marker, in HCC cells following LPS treatment and PAK1IP1 knockdown. Remarkably, PAK1IP1 knockdown led to increased IL-1β expression in HCC cells. Flow cytometry demonstrated an elevated apoptosis rate in LPS-treated HCC cells with PAK1IP1 knockdown. Additionally, WB revealed a significant upregulation of CASP-3 in response to PAK1IP1 inhibition. Subsequent experiments involved si-PAK1IP1 knockdown and treatment with Z-DEVD-FMK, a CASP-3 inhibitor, to evaluate the influence on the proliferation, invasion, and migration of HCC cells. Notably, PAK1IP1 knockdown inhibited the activities of LPS-treated HCC cells, while Z-DEVD-FMK treatment promoted these cellular processes. Collectively, our experimental findings demonstrate PAK1IP1 regulated the level of pyroptosis-related genes via the CASP-3-dependent pyroptosis.

In this study, we proposed a potential role for PAK1IP1 in hepatocellular carcinoma and explored its correlation with prognosis and immune cell infiltration. However, we must recognize that some aspects of this study have limitations. First, although we validated the function of PAK1IP1 by qRT-PCR, WB and cellular experiments, these experiments were mainly limited to cell lines. We have not yet assessed the relevance of these findings in the *in vivo* tumor microenvironment, which may limit the clinical applicability of our conclusions. Second, regarding apoptosis studies, we focused on CASP-1/CASP-4 and GSDMD cleavage, but failed to examine GSDME cleavage, which may play a role in CASP-3-dependent inflammatory apoptosis. Furthermore, although our study proposed a risk model containing 26 prognostic genes, external validation in an independent dataset is needed to assess the generalizability and reliability of the model. Regarding functional enrichment analysis and pathway identification, we recognized that only limited experimental validation has been performed on the turquoise module. Future studies will require more comprehensive experimental validation of the functional roles of these genes in liver cancer biology. More extensive *in vivo* studies, including additional markers of inflammatory apoptosis, as well as external validation of our prognostic models would be valuable next steps when considering future research directions. In addition, in-depth studies of the broader role of PAK1IP1 in hepatocellular carcinoma biology are essential. This study revealed a possible important role of PAK1IP1 and other factors in hepatocellular carcinoma progression and patient prognosis, but further experimental and clinical studies are needed to deepen our understanding and confirm these preliminary findings.

## Conclusions

To sum up, PAK1IP1 functions as a proto-oncogene in HCC. Additionally, the increased infiltration of myeloid dendritic cells observed in HCC samples highlights their potential as immunological targets. Furthermore, our *in vitro* experiments elucidated the mechanism of PAK1IP1, revealing that PAK1IP1 knockdown induced CASP-3-dependent pyroptosis in HCC cells to inhibit HCC progression. These findings laid the groundwork for future research endeavors for developing new clinical biomarkers.

## Data Availability

All relevant data are within the manuscript and its Supporting Information files.

## Availability of data and materials

The datasets used and/or analyzed during the current study are available from the corresponding author on reasonable request.

## Ethics approval and consent to participate

Not applicable.

## Patient consent for publication

Not applicable.

## Author Contributionthe

1. Conception and design of the study, or acquisition of data, or analysis and interpretation of data: Xiaoliang Lu and Jie Chen
2. Drafting the article or revising it critically for important intellectual content: Jie Chen and Hong Zang
3. Final approval of the version to be submitted: Zefa Lu and Xiaoliang Lu

## Acknowledgments

We express our deepest gratitude to all those who have made this study possible. Our heartfelt thanks go to the team members for their collaboration and diligent work, and to them who provided insight and expertise that greatly assisted the research. All authors have read and confirmed the acknowledgment.

